# Isolation Considered Epidemiological Model for the Prediction of COVID-19 Trend in Tokyo, Japan

**DOI:** 10.1101/2020.07.31.20165829

**Authors:** Motoaki Utamura, Makoto Koizumi, Seiichi Kirikami

## Abstract

**Background:** Coronavirus Disease 2019 (COVID19) currently poses a global public health threat. Although no exception, Tokyo, Japan was affected at first by only a small epidemic. Medical collapse nevertheless nearly happened because no predictive method existed for counting patients. A standard SIR epidemiological model and its derivatives predict susceptible, infectious, and removed (recovered/deaths) cases but ignore isolation of confirmed cases. Predicting COVID19 trends with hospitalized and infectious people in field separately is important to prepare beds and develop quarantine strategies.

**Methods:** Time-series COVID19 data from February 28 to May 23, 2020 in Tokyo were adopted for this study. A novel epidemiological model based on delay differential equation was proposed. The model can evaluate patients in hospitals and infectious cases in the field. Various data such as daily new cases, cumulative infections, patients in hospital, and PCR test positivity ratios were used to examine the model. This approach derived an alternative formulation equivalent to the standard SIR model. Its results were compared quantitatively with those of the present isolation model.

**Results:** The basic reproductive number, inferred as 2.30, is a dimensionless parameter composed of modeling parameters. Effects of intervention to mitigate the epidemic spread were assessed a posteriori. An exit policy of how and when to release a statement of emergency was also assessed using the model. Furthermore, results suggest that the rapid isolation of infectious cases has a large potential to effectively mitigate the spread of infection and restores social and economic activities safely.

**Conclusions:** A novel mathematical model was proposed and examined using COVID19 data for Tokyo. Results show that shortening the period from infection to hospitalization is effective against outbreak without rigorous public health intervention and control. Faster and precise case cluster detection and wider and quicker introduction of testing measures are strongly recommended.

## INTRODUCTION

Coronavirus Disease 2019 (COVID-19) currently represents a global public health threat. Tokyo, Japan is no exception, but its epidemic was small despite lacking rigorous public health intervention. The thorough behavior changes of individuals, with social distancing, avoidance of closed spaces with poor ventilation, crowded places with many people, and close-contact settings such as close-range conversation (3 Cs) appear to explain Japan’s ability to slow the spread of COVID-19. Medical collapse, however, nearly occurred because of the lack of beds to accommodate the increasing number of patients. Therefore, to cope with future epidemics, mathematical prediction tools are thought to be indispensable to anticipate the maximum number of patients under treatment.

From a clinical perspective, COVID-19 has a long incubation period. Moreover, it is infectious even before the onset of symptoms. We recognize the presence of infection through testing, but only with delayed timing, which might exacerbate its spread. Measurements can express the appearance of past infections. Therefore, real-time conditions cannot be known from future measurements. To take proper preventive action, some mathematical model is necessary to infer present conditions. The standard Susceptible–Infectious–Removed (SIR) epidemic model [1] and most of its modified versions [2,3,4] have three components, with numbers of people who are susceptible, infected, and removed. Nevertheless, they do not include consideration of isolation of infected individuals. A four-component susceptible, exposed, infected, and removed (SEIR) model, [5,6] and its delayed model [7,8] have also been assessed. Many works reported to date have specifically examined basic reproductive numbers for COVID 19. Khalid Hattaf et al. reported that including incubation periods lowers the basic reproductive number [5].

Preparing the necessary number of beds, developing countermeasures, and scheduling their use necessitates development of a model to predict COVID-19 trends of hospitalization as well as infectious cases in the field separately. Nevertheless, few reports to date have considered the isolation of confirmed cases [4,16]. Shao et al. [4] predicted a future scenario of epidemic evolution in Tokyo using cumulative confirmed data from January 20, 2020 through February 21, 2020, which caused overestimation of cumulative infections and unfortunately yielded little information related to hospitalized cases.

This paper proposes a new delay differential equation with a primary unknown variable chosen as cumulative confirmed cases including isolation (hospitalized). The present epidemic model was examined through various time series data obtained in Tokyo [15]. The relation between the fundamental reproduction number R0 and the parameter of the present model was discussed. Differences in calculations between the standard SIR epidemic model and the present one were quantified. Furthermore, based on this knowledge, an exit strategy of the first epidemic and an inlet strategy for the second epidemic were assessed.

## METHODS

### Data

For this study, we used a publicly available dataset of COVID-19 provided by the public health authority of Tokyo, Japan. The present epidemiological model was verified through various time series data from February 28, 2020 through May 23, 2020, up to two days before the ‘statement of emergency’ was issued by the government of Japan. The model includes cumulative confirmed infectious cases, daily new cases, confirmed and hospitalized, the number of patients under treatment, and recovered/deaths in hospitals. Results confirmed a positivity-ratio in PCR tests in Tokyo [15]. These data were collected by public authority announcements. Therefore, ethical approval is not required for the present study.

### Prediction Models

#### Standard SIR Epidemiological Model and Its Alternative Formulation

The standard SIR epidemiological model is formulated as

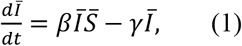

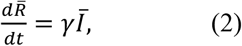

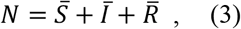

where variables 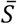, 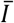, and 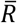 respectively denote susceptible, infectious, and removed and *N* represents the collective population under consideration. Also, β and γ are the parameters to be determined. Here, we introduce cumulative infections *x* as the primary unknown variable. A delayed differential equation equivalent to Eqs. (1)–(3) can be obtained as follows. Substituting Eq. (2) into Eq. (1) and noting the relation 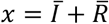 Eq. (1) can be rewritten as

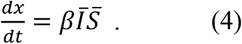

Differential operator *d*/*dt* represents the first-order delay with time constant 1/γ; it is equivalent to substitute 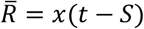 for Eq. (2) in which all infectious cases were assumed to be removed after a period of time *S*. Then, with 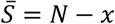 from Eq. (3), one finally obtains a newly found delay differential equation with a single unknown variable *x* equivalent to Eqs. (1)–(3) as

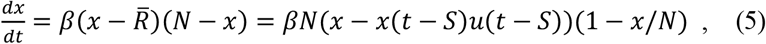

where *u* is the step function defined as

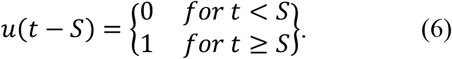

In this formulation, input parameter *S* can be ascertained from measurements. We set *S* as representing the sum of time interval *T* and treatment days in hospital until discharge. The latter was referred from official data of Tokyo [15].

#### Epidemiological Model Considering Isolation

The concept of the present model is presented in **Fig. 1**. In this epidemiological model considering isolation, cumulative infections *x* represent the number of onset of clinical symptoms among cases of infection and do not include asymptomatic infections (silent spreaders). Time interval *T* represents the period of time from infection until hospitalization. It is the sum of the incubation time and detection and testing times. WHO announced that the incubation time of COVID-19 was 7 days [10]. In other reports of the literature, it was reported as 5–6 days [8,9,12].

**Figure 1.**
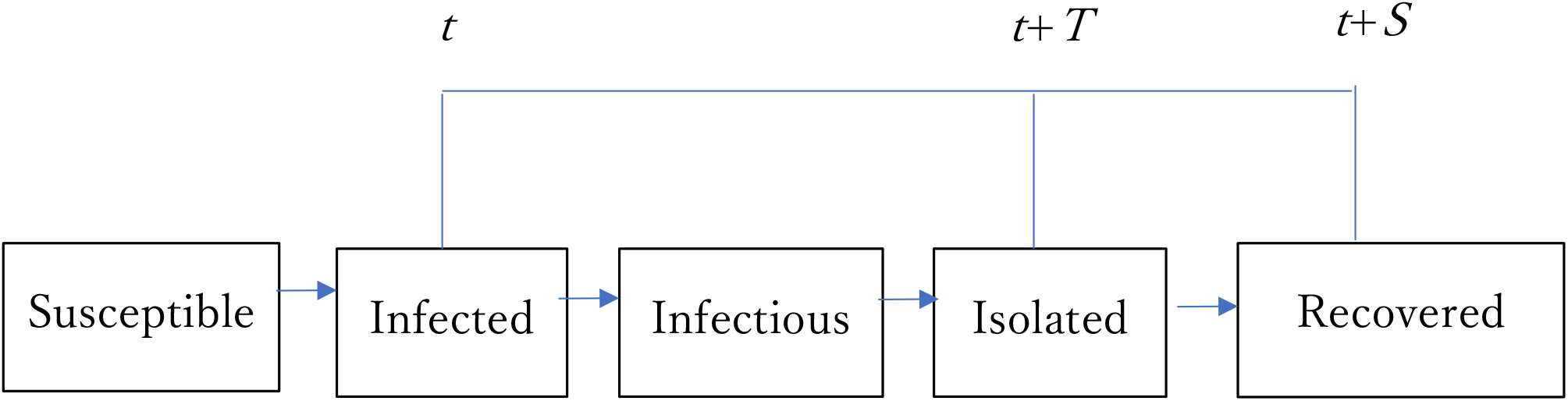
Concept of the present epidemic model.

For simplicity, the following assumptions were made.

1. Those cases infected before *t*-*T* are all isolated inside a ward denoted by *Y*, which can be expressed as *x*(*t*-*T*). Those infected in period [*t*-*T*, *t*] denoted by *x*-*Y* (=Q) exist outside the ward.
2. We assumed Q are infectious from infected until hospitalized and contain no removed (recovered/deaths) because T<S. As removal happens solely in hospital, the recovered cases and deaths in hospitals are *Z*. Patients under treatment in hospitals denoted by *P* can be represented as *Y*-*Z*. Therefore, *P*+*Q* becomes *x*-*Z* and corresponds to 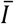 in the SIR model.
3. The spread of infections inside wards was not examined in the present model. Consequently, in the present model, 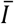 in Eq. (4) was replaced by *Q* in the present model. Then, we have a governing delay differential equation with a single unknown variable *x* as

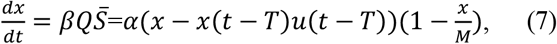

where α(=βM) is the transmission rate. We will designate the above-described formulation as the Apparent Time Lag Model (ATLM) hereinafter.

Here, α depends on the personal behavior of individuals in daily life such as social distancing, washing hands, masking, public health intervention, and control. *M* represents potential susceptible people at the inlet to an epidemic. *M* is not a geographical population, but a virtual one. In fact, *M* is much smaller than the actual population of Tokyo because the daily action range of a normal person is limited where the possible number of people that a person would meet is not large. Therefore, *M* cannot be ascertained before calculation: instead, it is inferred from parameter fitting to measurement data. Once Eq. (7) is solved, important infectious variables are derived in terms of *x*.

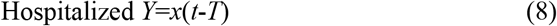

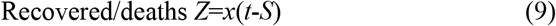

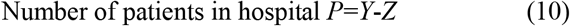

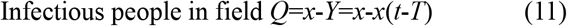

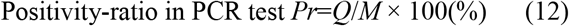

**Table 1** presents the results.

**Table 1.** Expressions of variables.

| Table 1 Expressions of variables. |  |  |
| --- | --- | --- |
| Item | Variable | expression |
| Cumulative onset | $x$ | |
| Hospitalized | $Y$ | $x(t-T)$ |
| Recovered/deaths | $Z$ | $x(t-S)$ |
| Patients in hospital | $P$ | $Y-Z$ |
| Infectious in field | $Q$ | $x-x(t-T)$ |
| Positivity-ratio in PCR test | $Pr$ | $Q/M^*100(\%)$ |

Eq. (7) can be reduced to dependence on the respective time spans.

For 0<*t*<*T*,

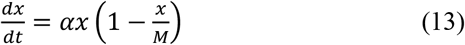

,where infectors in this period are expressed by *x*. The first term of RHS is governing Eq. (13) for *x*<*M*/2; so is the second term for *x*>*M*/2. Then, *dx*/*dt* has a maximum at *x*=*M*/2.

This equation has an analytic solution, a so-called logistic function, as

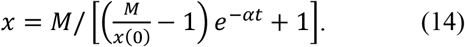

At the beginning, Eq. (14) shows an exponential epidemic growth yielding

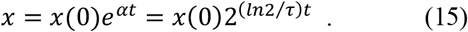

From this, α is related to clinical doubling time τ, as shown below.

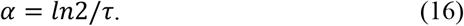

For example, with α being 0.164, the equivalent value of τ would be 4.22 days; *x*/*x*(0) would become 10 in two weeks. Now, parameter α has a clear physical meaning.

For *t* > *T*,

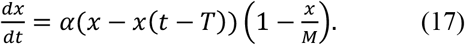

In the case of 1≫*x*/*M* Eq. (17) reduces further to

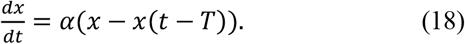

Eq. (18) has an approximate analytic solution of the form (see **Appendix A**)

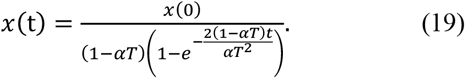

The trend of *x*(*t*) is governed by a dimensionless parameter α*T*, which exhibits threshold of bifurcation because of nonlinearity nature of time delay term *x*(*t*-*T*) in Eq. (18). If it is larger than unity, *x*(*t*) would increase exponentially (outbreak). Otherwise it would converge to *x*(0)/(1-α*T*) (endemic). At the boundary where α*T* approaches unity, *x*(*t*) would increase linearly such that

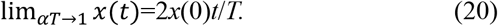

Therefore, it is noteworthy that α*T* characterizes the trend at an early stage of the epidemic.

#### Consideration of Silent Spreader

We will extend the model above to accommodate silent spreaders to evaluate antibody production. Silent spreaders are asymptomatic but infectious people. Therefore, no isolation is possible for them. For simplicity we assume they continue to be infectious from infected until recovered. Presumably, the percentage of antibodies of silent spreaders is known a priori. It is independent of the model parameters described above. Total infectious cases in the field are presumed to be the sum of cases from silent spreaders and from that with onset of symptoms that obeys Eq. (7). Let ε be the share of silent spreaders, infectious term *Q* in Eq.(7) might be replaced by (1-ε)*Q* + ε(*x*-*Z*) in this case. Consequently, we have a differential equation for the model including silent spreaders as

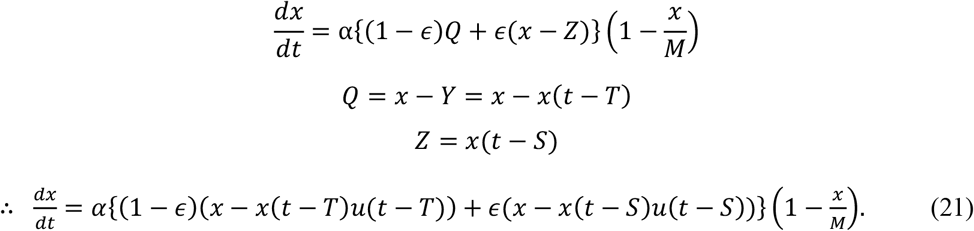

### Reproductive number

The effective reproductive number *Rt* implies the number of infections per single representative infection in a collective population until it is removed. Various calculation methods have been reported [5,6,8,9] for *Rt*. In this model, possible infectors affecting their infectees at time *t* exist in a time span [*t*-*T*, *t*] because the infectors produced before *t*-*T* are all hospitalized. Consequently, for this paper, *Rt* was calculated as the ratio of daily new cases to the average number of infectants per day during time span [*t*-*T*, *t*] as

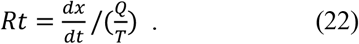

Combination of Eq. (22) with Eq. (7) yields

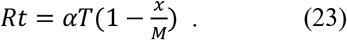

Because no preventive measure is taken at an initial stage of first epidemic, *Rt* at the initial phase coincides with basic reproductive number *R*0. Then, noting 1≫*x*(0)/*M*, we have

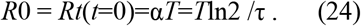

Dimensionless parameter α*T* was identified as the basic reproductive number R0 in the present model. Furthermore, the physical meaning of R0 was clarified, which is the product of the ratio of incubation time and epidemic doubling time multiplied by ln2. However, Makino reported that the standard SIR epidemic model expressed by Eqs. (1)–(3) provides *βN*/*γ* for the basic reproductive number [17].

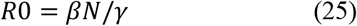

Recalling *βN*=*α* and 1/*γ* as the time constant, Eq. (25) has a meaning similar to that of Eq. (24).

For numerical integration of Eqs. (5), (7), and (21), 4th order Runge–Kutta–Gill (RKG) method was applied in the following manner. To solve the following equation of 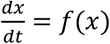, the RKG scheme becomes

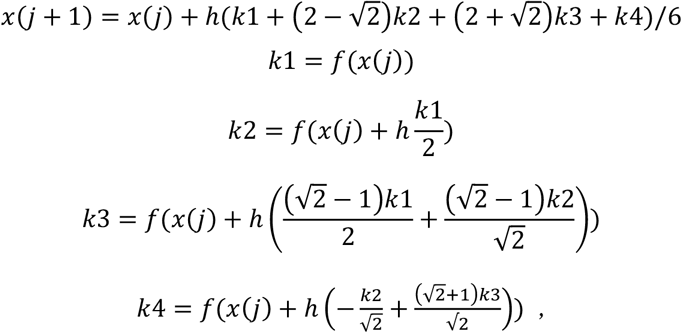

where *h* stands for the time step. With *h* chosen as one day, accuracy was found to be inadequate for evaluating epidemic cases. Consequently, *h* was chosen to represent half a day.

## RESULTS

### Accuracy of the Numerical Integration Scheme

**Figure 2** presents a comparison of the analytic solution (Eq. (14)) with numerical integration of Eq. (13) using RKG method. Parameter values were *x*(0)=3, *M*=6200, and α=0.164. Time step *h* was chosen as half a day. Excellent agreement is obtained for both cumulative and daily new cases. Relative error for *x* at *x*=*M*/2 where the curve becomes steepest was confirmed to be 8*10^(−7) that is adequately small. Then the RKG scheme with *h*=0.5 was adopted for the following analyses.

**Figure 2.**
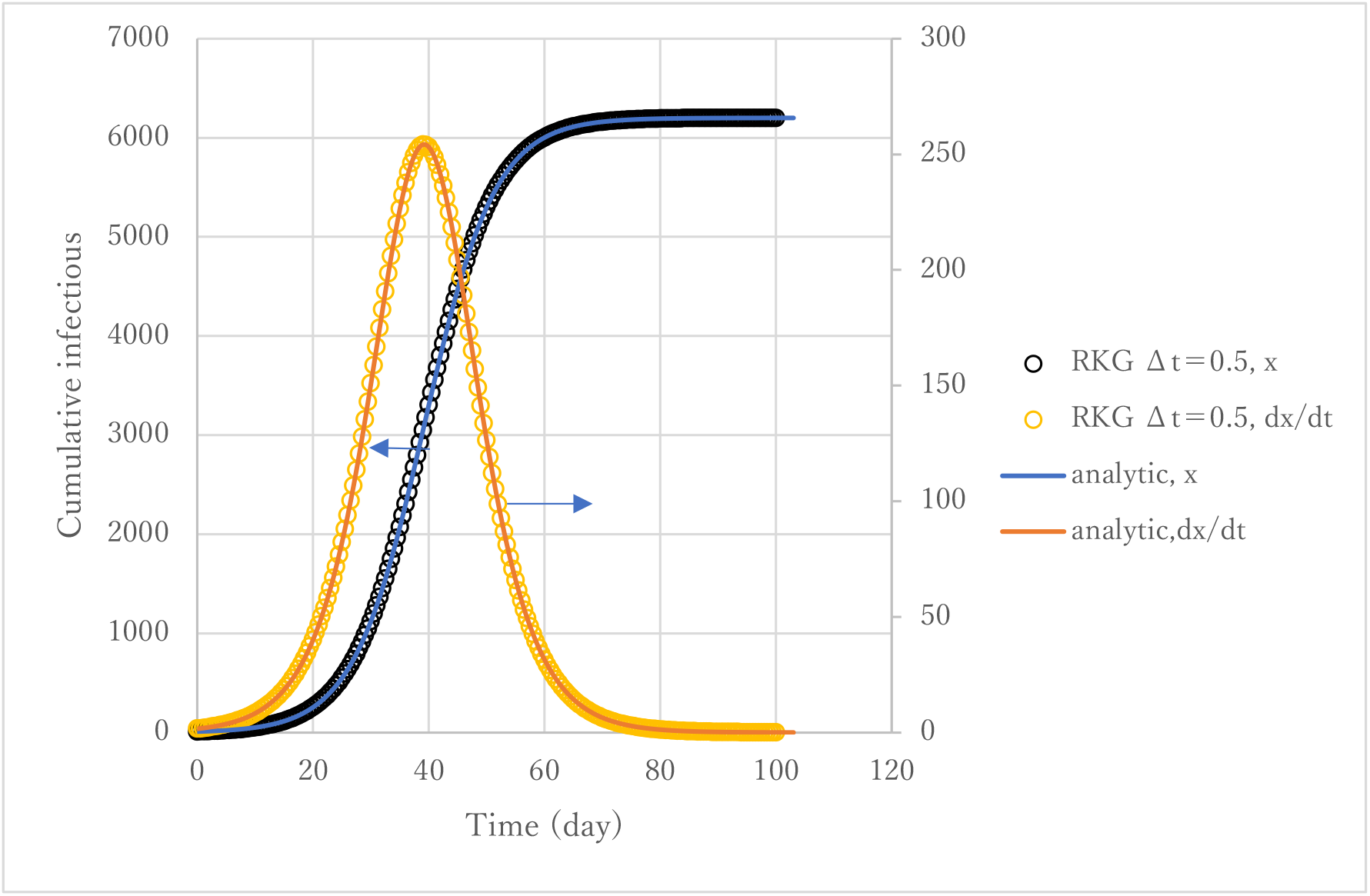
Accuracy of numerical integration scheme.

### Dimensionless Parameter to Characterize Early Stage Epidemic Trends

**Figure 3** presents an early stage epidemic trend of the solution *x*(*t*) of Eq. (7). Clear change in the behavior of *x*(*t*) depending on the value of α*T*, so-called bifurcation nature, is apparent.

**Figure 3.**
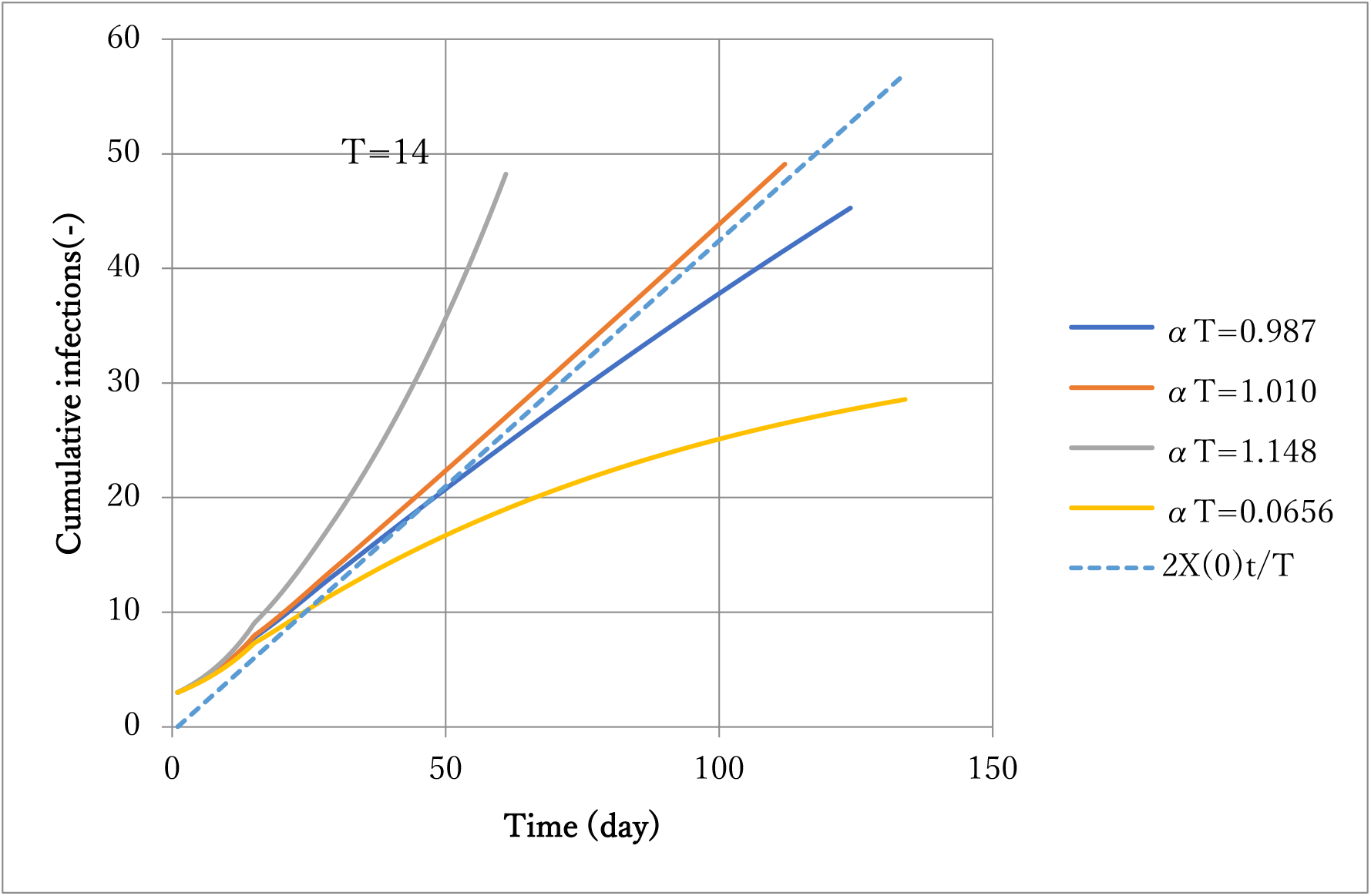
Effect of parameter *αT* on solution *x*.

Actually, α*T*=1 is found to be a threshold with α*T*>1 causing an outbreak and with α*T*<1 as endemic. This fact is consistent to the interpretation that α*T* is equal to R0 in Eqs. (22)-(24) aforementioned.

### Comparison of ATLM Prediction with Observed Data

Cumulative confirmed cases of February 28 – April 28 in the first epidemic of COVID-19 in Tokyo were applied for the fitting of parameters α and *M*. Values of other parameters *T* and *S* were preset as 14 and 36 based on empirical knowledge. The former has been widely acknowledged for COVID-19. The latter was derived as a sum of T and average stay days in hospital until discharge or death. According to parameter survey calculations, α and *M* were selected respectively as 0.164 and 6200. In the following, the number 0.164 will be designated as α0.

**Figure 4** presents a comparison of calculations against observed data in terms of cumulative hospitalized *Y*, the number of patients under treatment in hospital *P*, and removed (recovered and deaths) *Z* in hospitals. Whereas observed data are available for February 28 onward, calculations started with initial infection *x*(0)=3 on February 14, which was 14 days prior. The vertical line indicates the date April 28. Calculations succeeded in simulating observed data in general. Precisely, however, Y starts overestimating data in 77 days (May 2) and afterward. Probably, α became smaller because ‘Stay home’ was announced by metropolitan authorities five days earlier on April 27, by which people’s behavior changes lowered the value of transmission rate α.

**Figure 4.**
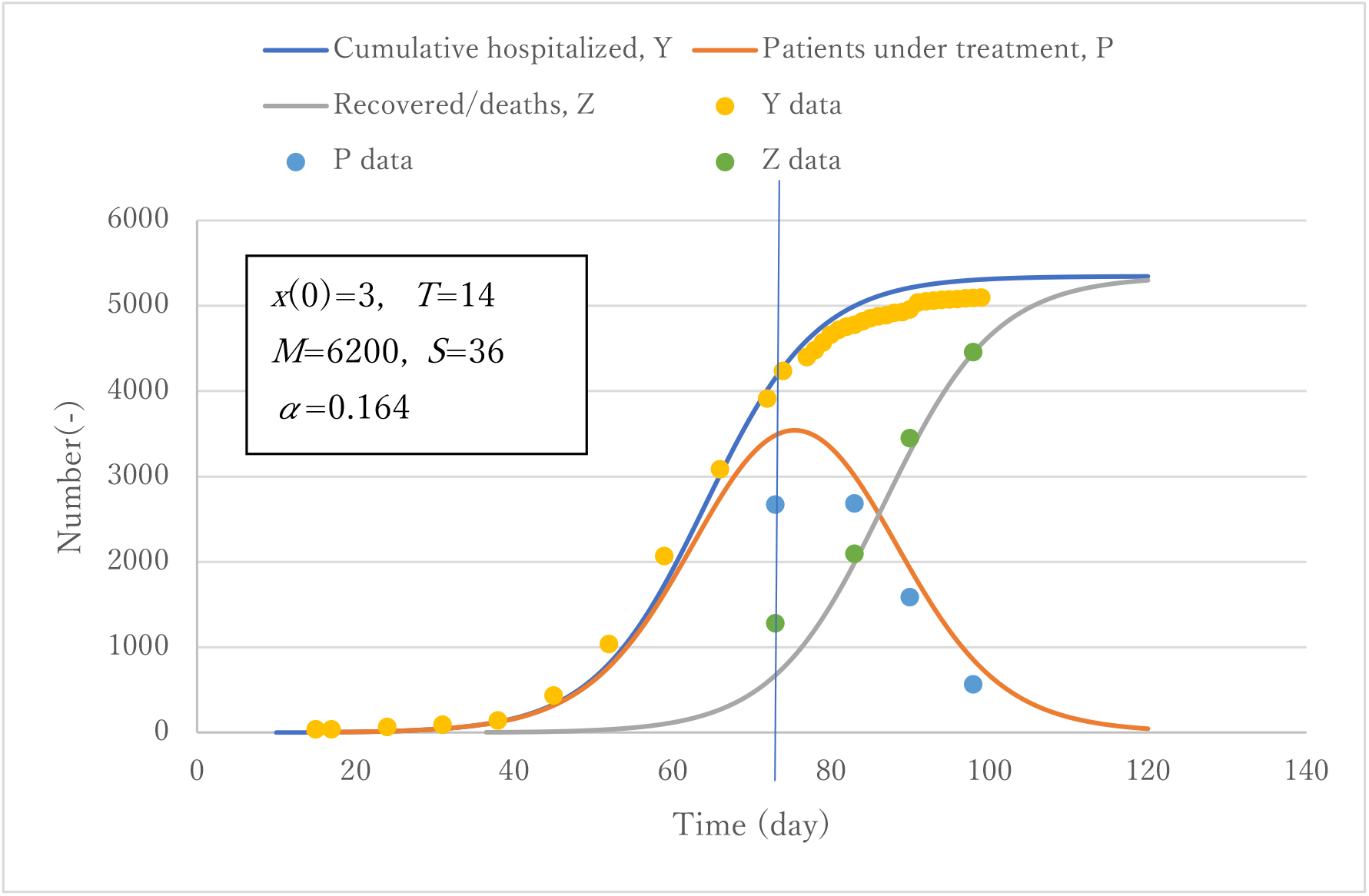
Verification of the model for hospitalized patients and removed.

**Figure 5** presents a comparison of prediction with observed data on daily new cases. Although the data show remarkable scattering, prediction can simulate their average trend at large. **Figure 6** exhibits observed trends of the positivity-ratio in PCR test compared with calculation *Pr* (Eq. (12)). In the case of Japan during the first epidemic, application of PCR tests was limited to those cases with onset of symptoms through a doctor’s examination in view of the tightness of the number of beds available. Data in the early stage might include large statistical error because of the fewer inspections conducted. Except for the period, however, data trends are seen to be well-simulated in spite of the model simplicity. That is true because the model counted infections with the onset of clinical symptoms, which suits the attribute of tested data [15]. This result proves that the present model (ATLM) can account for unconfirmed infectious cases outside ward that are not observable. The present ATLM allows implementation of public health intervention at the right magnitude and timing.

**Figure 5.**
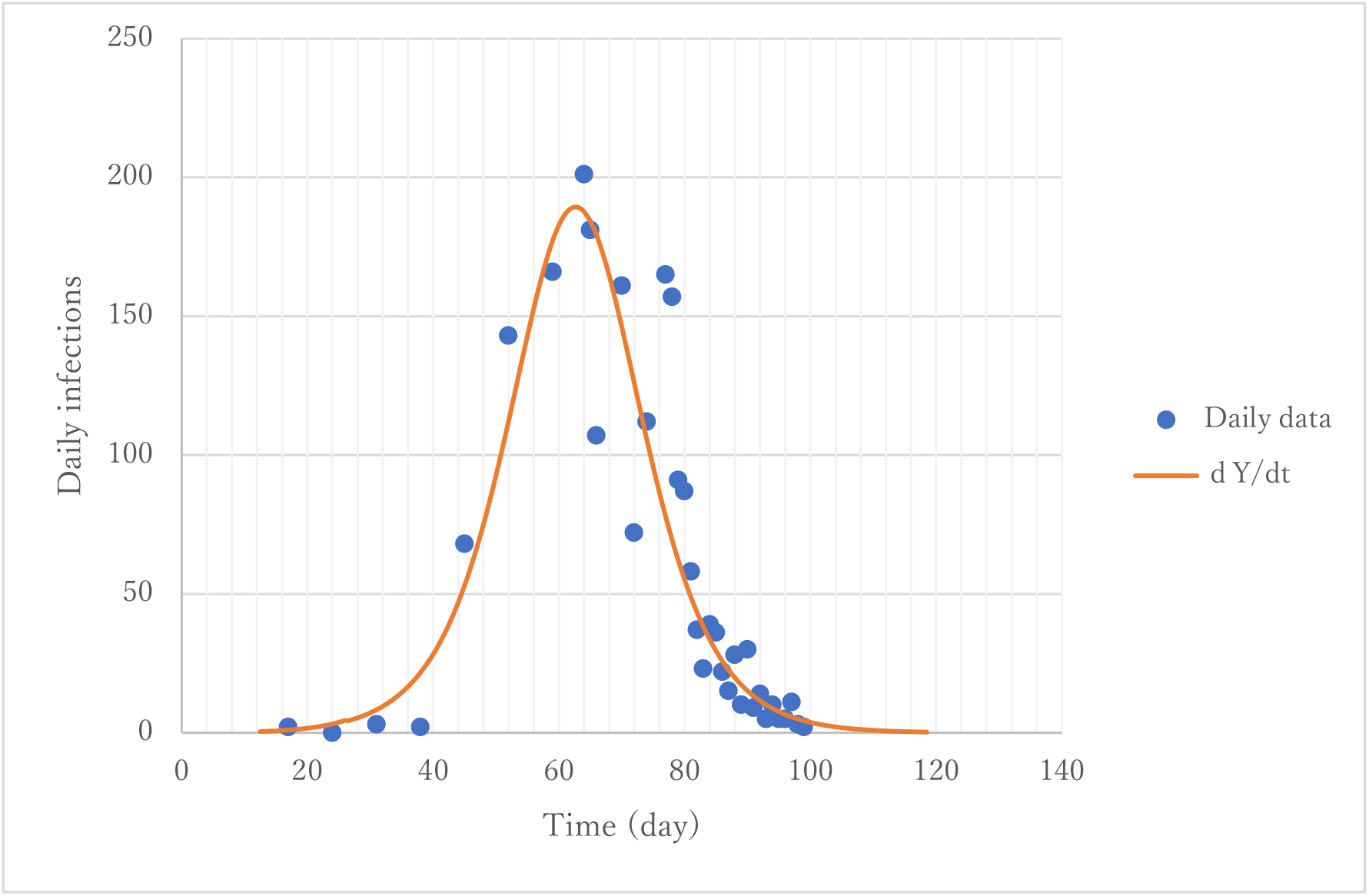
Simulation of daily new cases.

**Figure 6.**
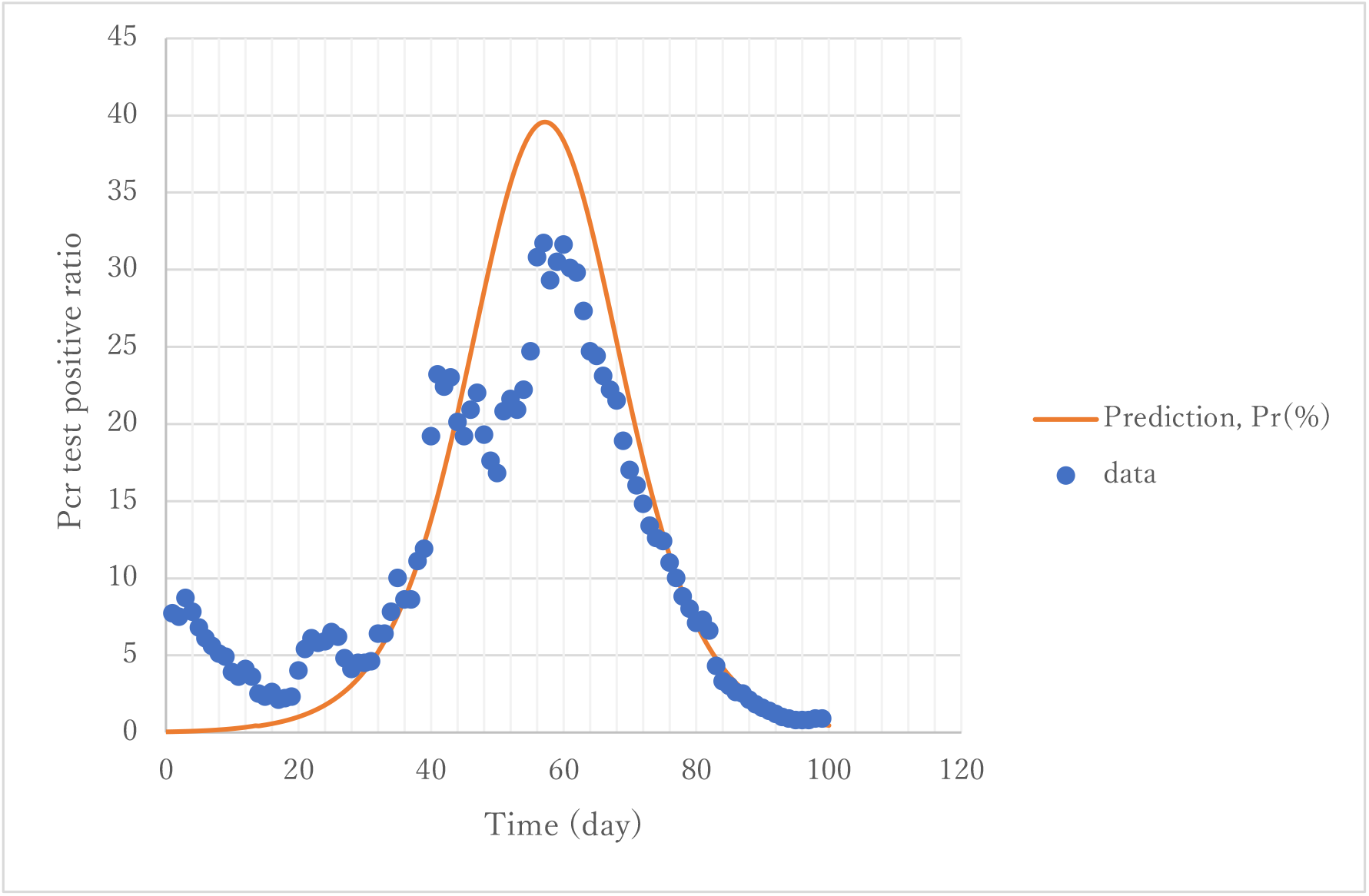
Simulation of positivity-ratio in PCR tests.

### Trend of Effective Reproductive Number

**Figure 7** gives a trend of field variables. Infectious cases in the field were predicted to have a peak of 2363 in 57 days (April 12), whereas the effective reproductive number Rt decreases monotonously to reach unity in 54 days (April 9). The vertical line indicates April 9 when it crosses the point where Rt=1. Both dates are close and reasonable. Susceptible persons remain uninfected by about 1000. The value of R0 is estimated as 2.30 using Eq. (24) with transmission rate α of α0. The value of α0, however, reflects the official statement of emergency issued on April 7. It might underestimate the value at the initial stage of an epidemic. Therefore, the actual R0 might be higher. Nonetheless, the value is close to the value 2.20 Li Qun obtained from the data taken in Wuhan, China [12] or 2.68 reported by Wu [14].

**Figure 7.**
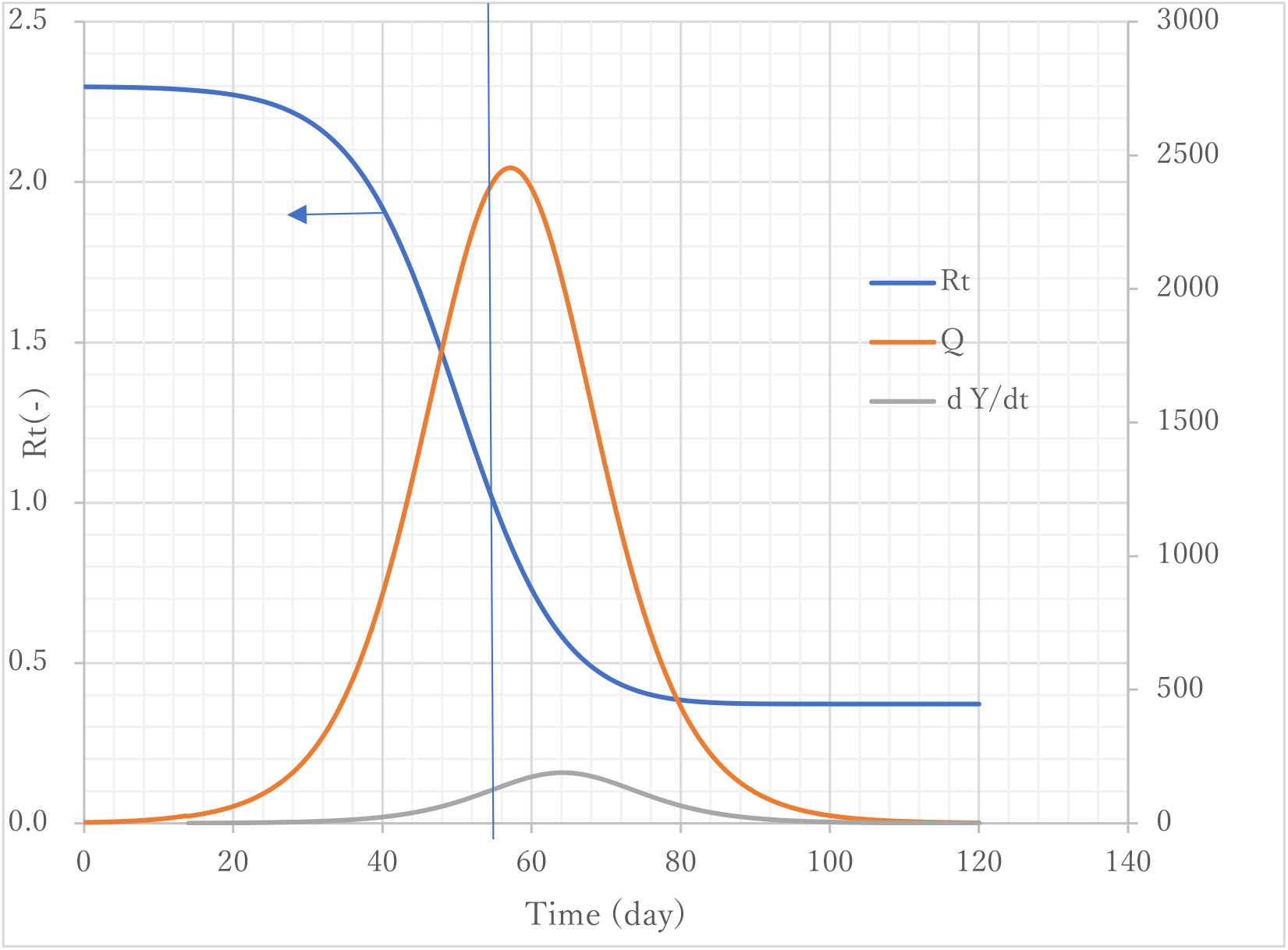
Trends of reproductive number and infectious cases in field.

### Comparison of ATLM with SIR

**Figure 8** presents a comparison of ATLM with standard SIR epidemiological models in terms of 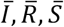 under the same parameter values. Their corresponding variables in ATLM are, respectively, *P*+*Q*, *Z*, and *M*-*Z*. The results obtainable by SIR are based on Eqs. (5) and (6), shown as a broken line and ATLM in the solid line. Marked differences are apparent between two. Actually, SIR predicted almost all the population as infected, although ATLM left behind 1000 population uninfected. Furthermore, ATLM gives lower values by 20% for infections, which results from modeling the hospitalization before removal.

**Figure 8.**
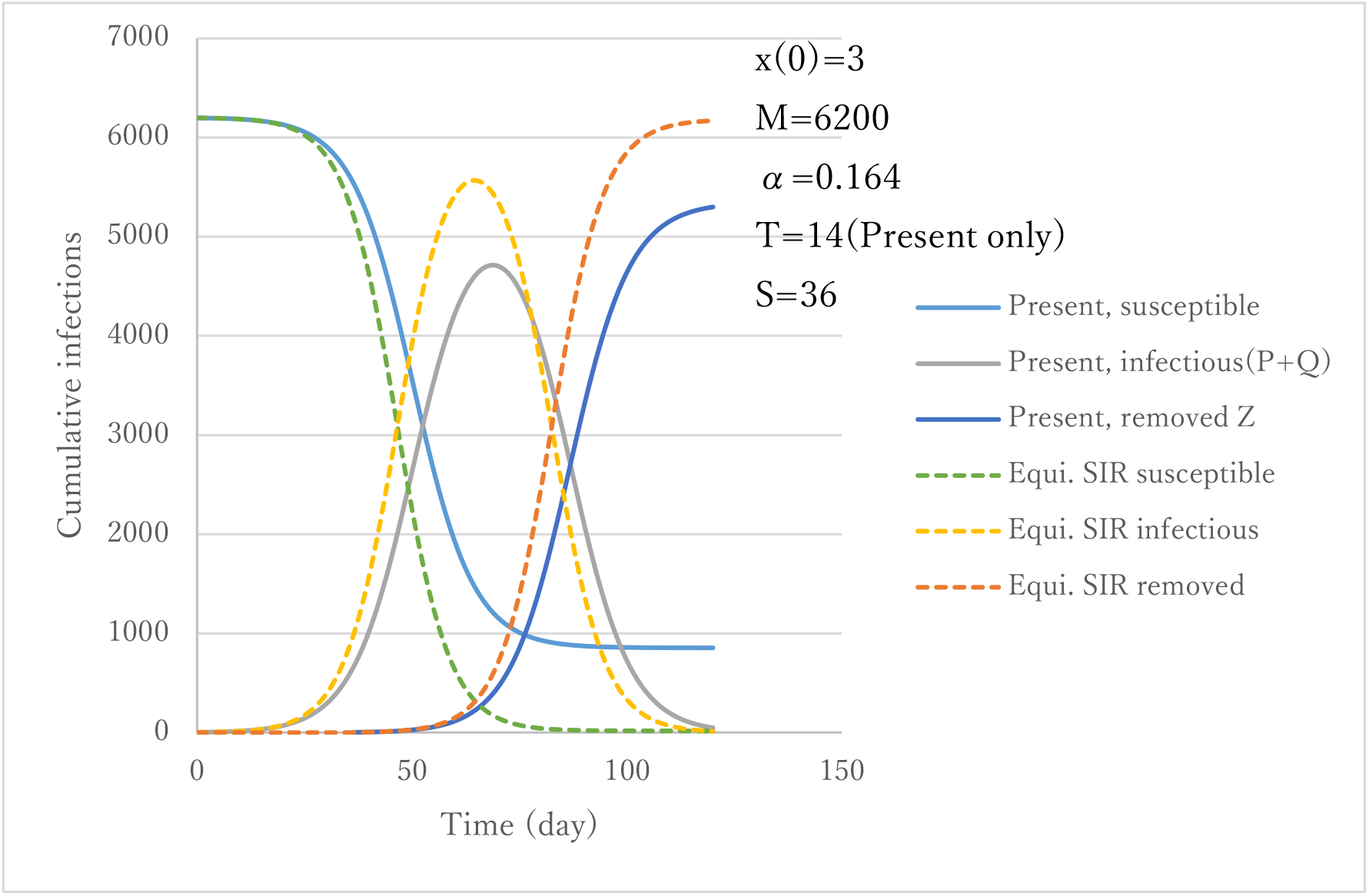
Comparison of a standard SIR model and the present ATLM model results.

### Population Balance of ATLM

**Figure 9** presents the concept of population balance in the present model. The outermost rectangle has an area of *M*: the collective population. The area of each compartment represents its cases. Initially frames 1 and 2 are on the right edge; with frame 3 on lower edge. As infection starts, frame 1 moves leftward, followed by frame 2 after elapsed time *S* when removal begins. Frame 3 moves upward after a period of time *T* when hospitalization begins. At the final stage of epidemic frame 2 catches up with frame 1 and both approach some horizontal location of the rectangle and frame 3 on the upper end. In this way, the sub-rectangle changes its area in proportion to their cases. **Figure 10** portrays a stacked graph with time under the conditions of *M*=6200, α=0.164, *T*=14, and *S*=36. The unit of the abscissa is half a day. Variables are located in order from bottom to top, with infected people in field *Q*, patients in hospital *P*, removed from hospitals *Z*, and susceptible *M*-*x*. Summation of all these values must coincide with the collective population *M* if the population balance is maintained. The value of top end is equal to 100% along the entire time span, which shows the balance at any instance maintained.

**Figure 9.**
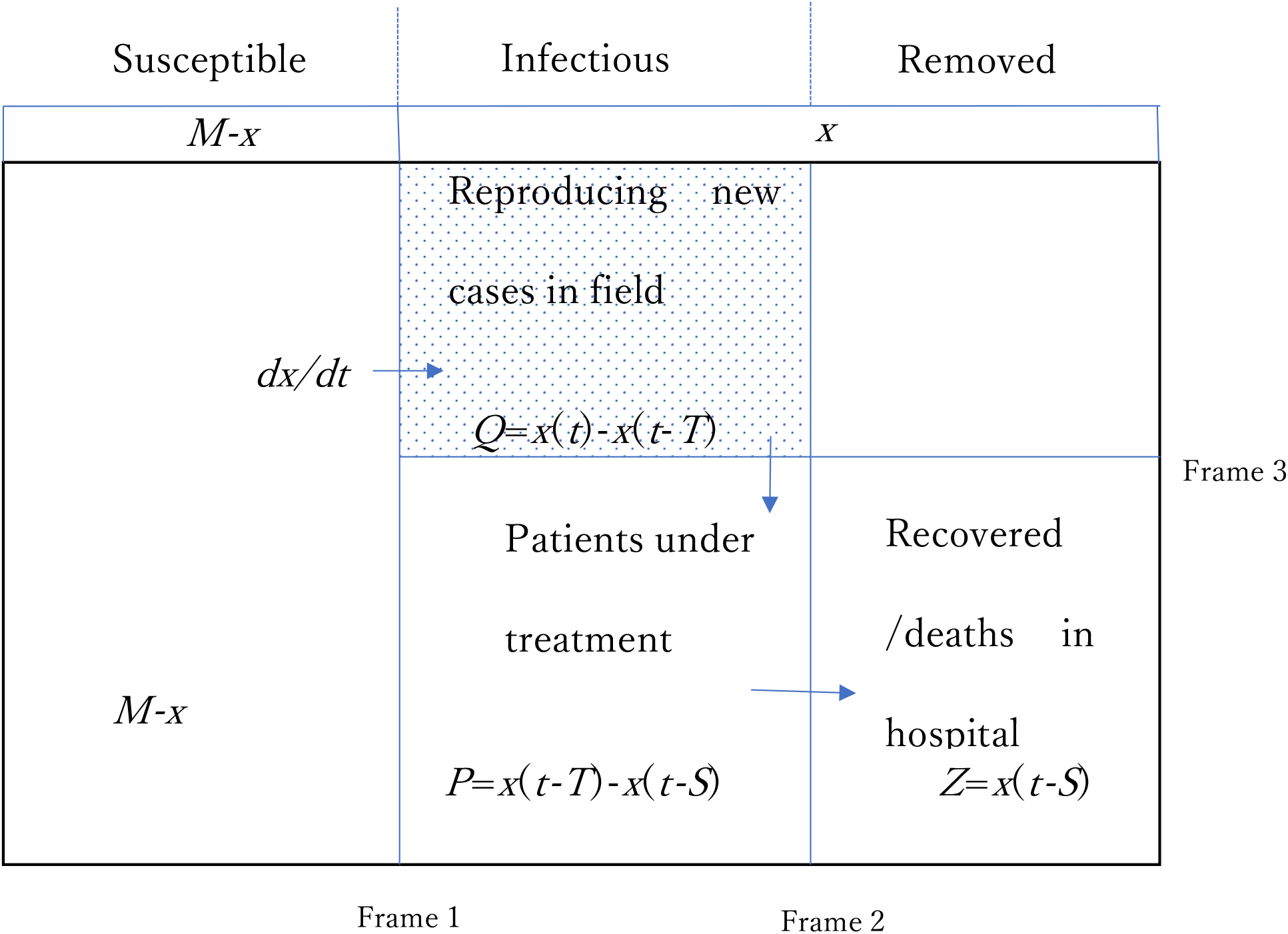
Concept of population balance of the present model.

**Figure 10.**
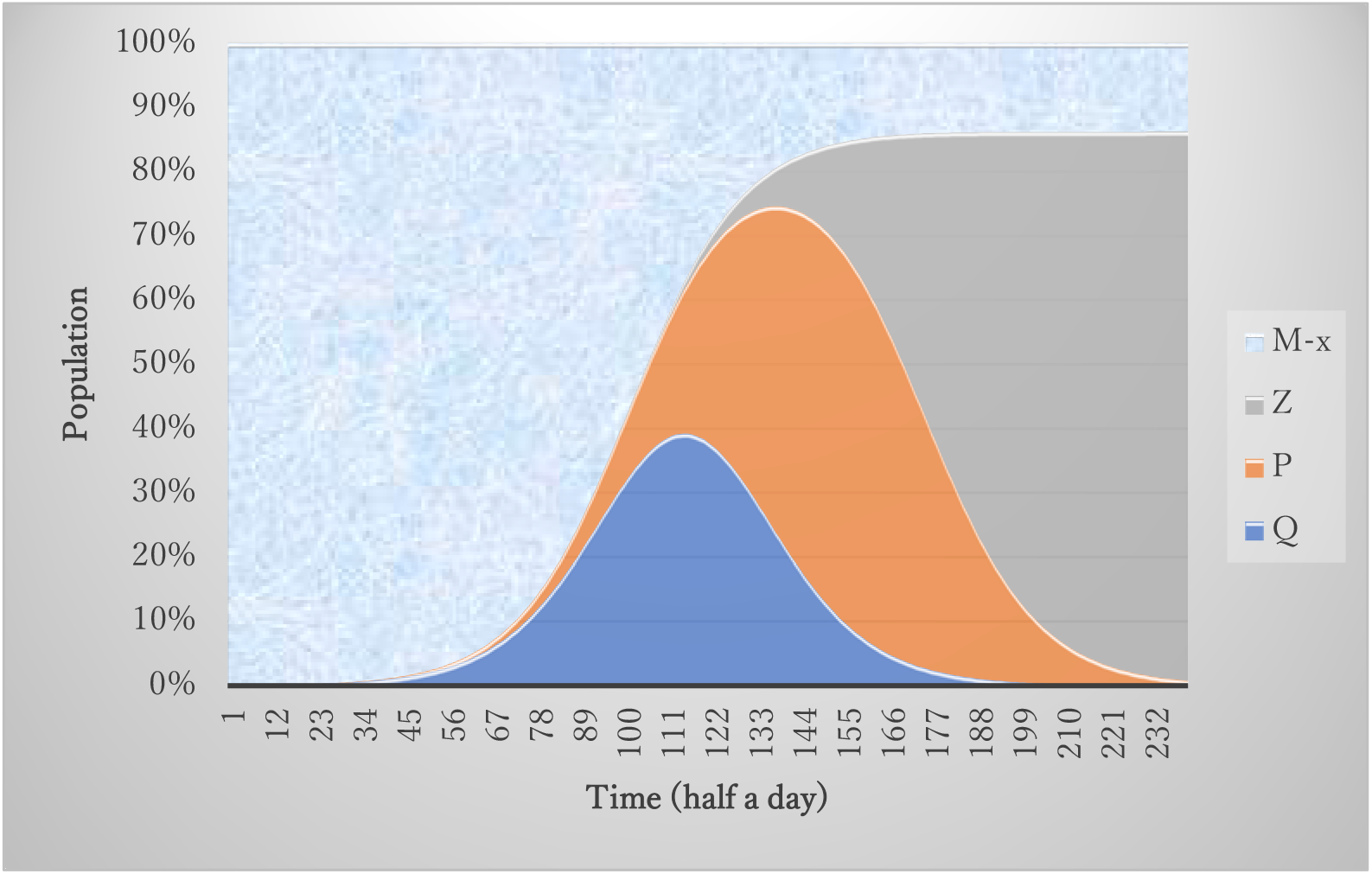
Population stacked graph by the present model.

### Antibody Production

According to the antibody tests conducted after the end of the first epidemic in Tokyo, its ratio was found to be 0.1%. Because the metropolitan population is 14 million, the number of individuals having antibodies is estimated as 14 thousand. Because the number of removed cases in the first epidemic was 5000, the rest are 9000 in the field. Taking ε 9000/14000=0.634 as the first guess, simulation of first epidemic was conducted using the antibody model to reproduce 5200 for (1-*ε*)*x*(∞) with change in ε. The best fit value of ε was 0.627. **Figure 11** presents a trend of various variables using Eq.(21) with silent spreaders. Compared with Q in **Fig. 7** the peak value of apparent spreaders inferred from antibody model is half in size. Apparent to silent spreaders ratio is 0.14 that is much less than the relative existence ratio (1-*ε*)/*ε*= 0.6. This may come from the difference of the values of T and S. As S/T is 2.6, a silent spreader can reproduce infectious cases more than an apparent spreader before removal. Further information on incubation as well as recovered periods of silent spreaders are needed for accurate modeling of antibody.

**Figure 11.**
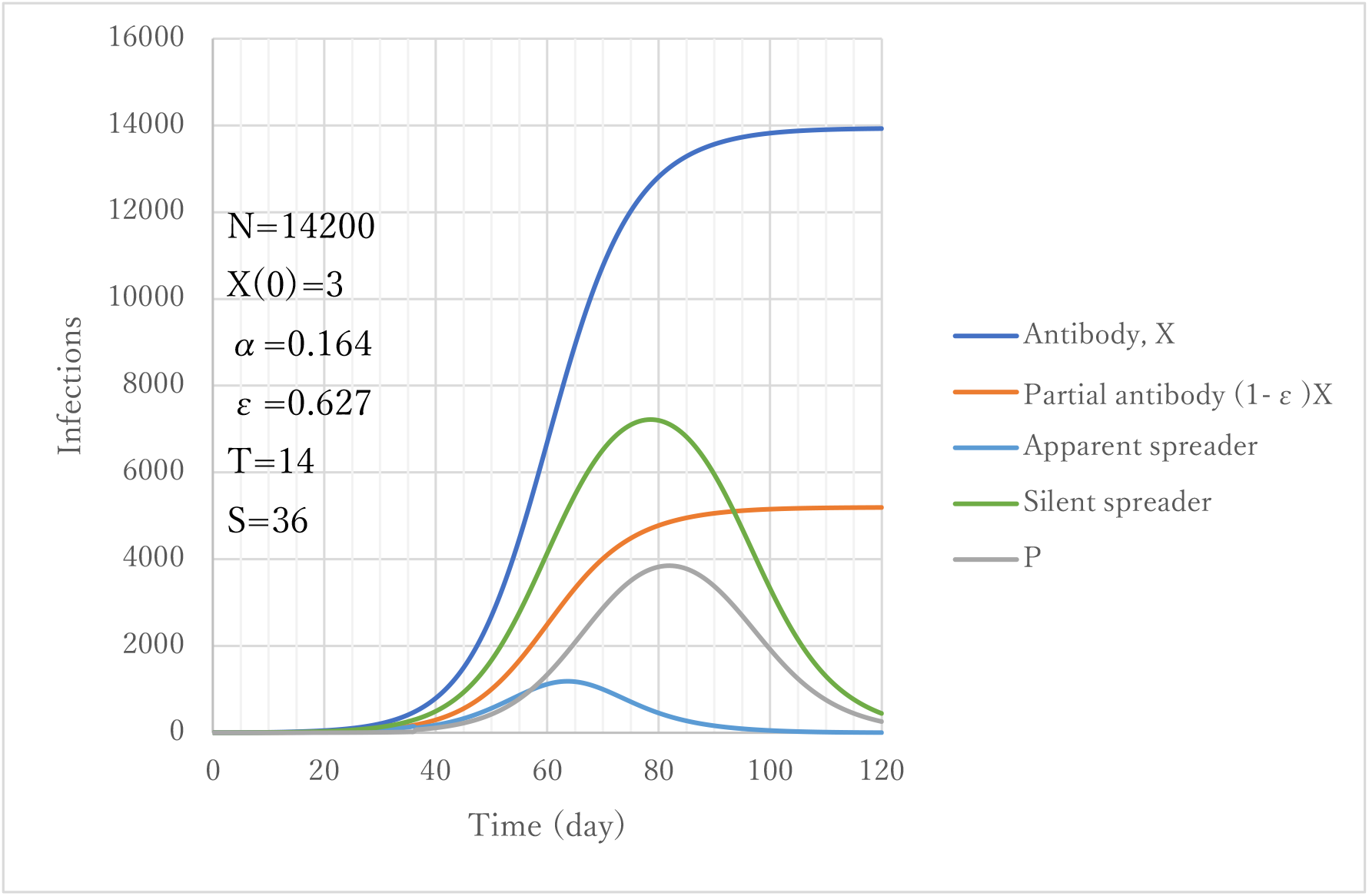
Model calculation with asymptomatic cases considered.

## DISCUSSION

### Intervention for Spread Mitigation

Toward the end of the first COVID-19 epidemic in Tokyo, strong public health intervention was conducted to reduce contact with people by 80%. The official announcement of ‘Stay home’ was declared at 72 days after onset of the epidemic. Simulation was made to assess its effects. Stepwise change in the value of parameter α from α0 to 0.2α0 or to 0.43α0 was applied at day 86 when the effect of the action should appear as delayed on daily new case data. The latter coincides with *Rt*=1. Actual reduction in the number of contacts was presumed to be 50–60%. Results are presented in **Fig. 12**. Three cases α0 (no action), 0.43α0 (Action B), and 0.2α0 (Action A) are given together with PCR test data. Unexpectedly, the data appear to follow the case of prediction of daily new confirmed cases without action *dY*/*dt*, which implies that the action failed. According to prediction by ATLM, the action should have been conducted earlier. In view of the trend of infectious cases in field Q calculated using ATLM, imaginary action with 0.43α0 (Action C) was taken at day 40, which appears on daily new cases data in two weeks later when computational action was made. A significant effect might have been obtained.

**Figure 12.**
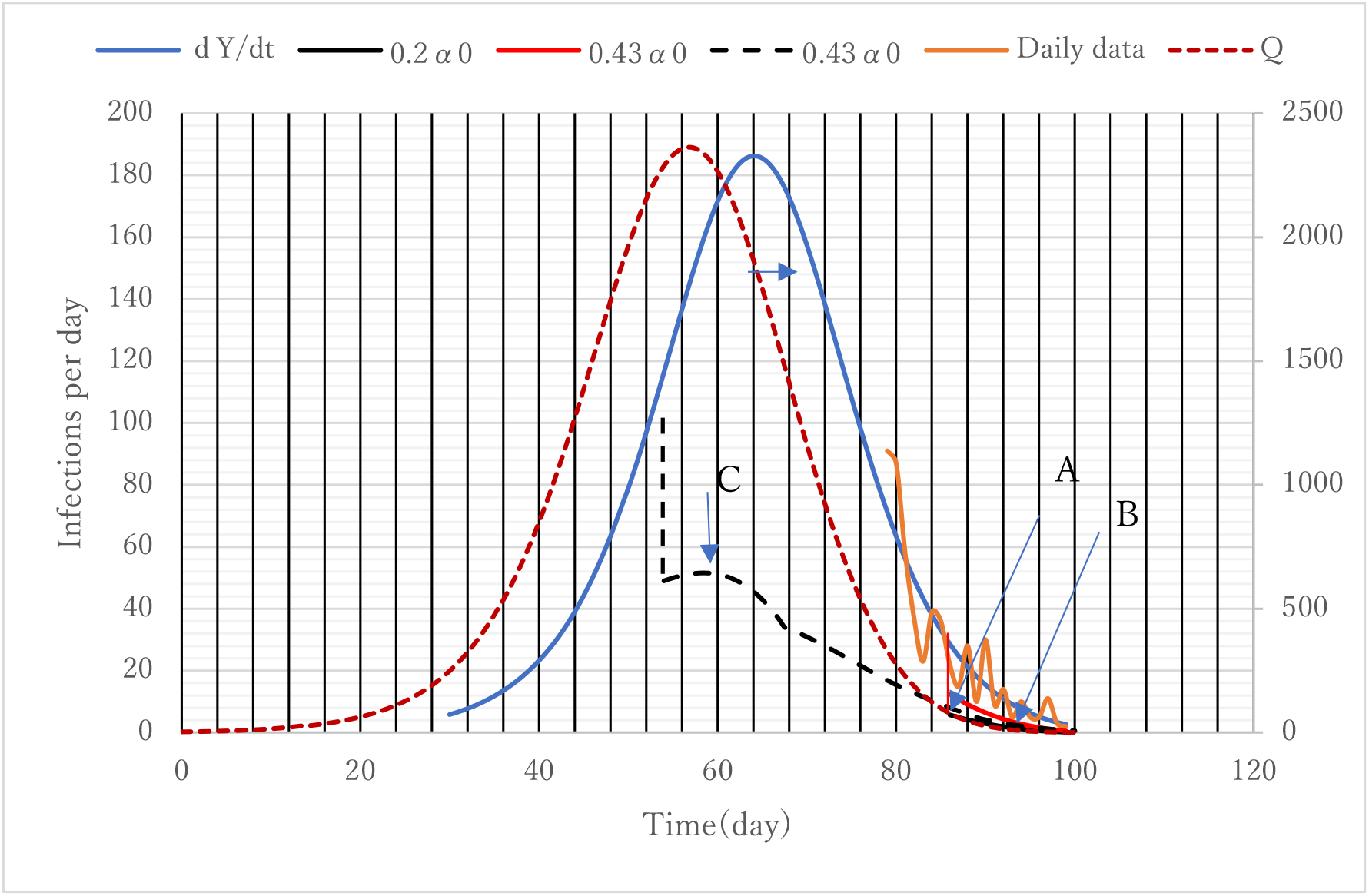
Quarantine strategy to reduce *α*.

### Exit Strategy for Endpoint

The Japanese government issued a criterion for rescission of the emergency statement to stay at home, stipulating that daily infections decrease to no more than 0.5 person/ day per population of 100 thousand. This, applied to Tokyo, the criterion would give 10 infections per day. Posterior assessment was made. **Figure 13** presents trends of infectious variables toward the end of the first epidemic in Tokyo. The criterion of 10 infections/day can be represented as *dY*/*dt* (broken line) at day 93 after onset of the first epidemic (February 14). Noting that measured values appear with a 14 day delay, the actual number is predicted to be 0.3 according to *dx*/*dt*. This number is below unity and is sufficiently low for rescission. Infectious cases in the field, however, are still around 10. An additional 7 days (100 days after) would be necessary to make it below unity to coronavirus extinction.

**Figure 13.**
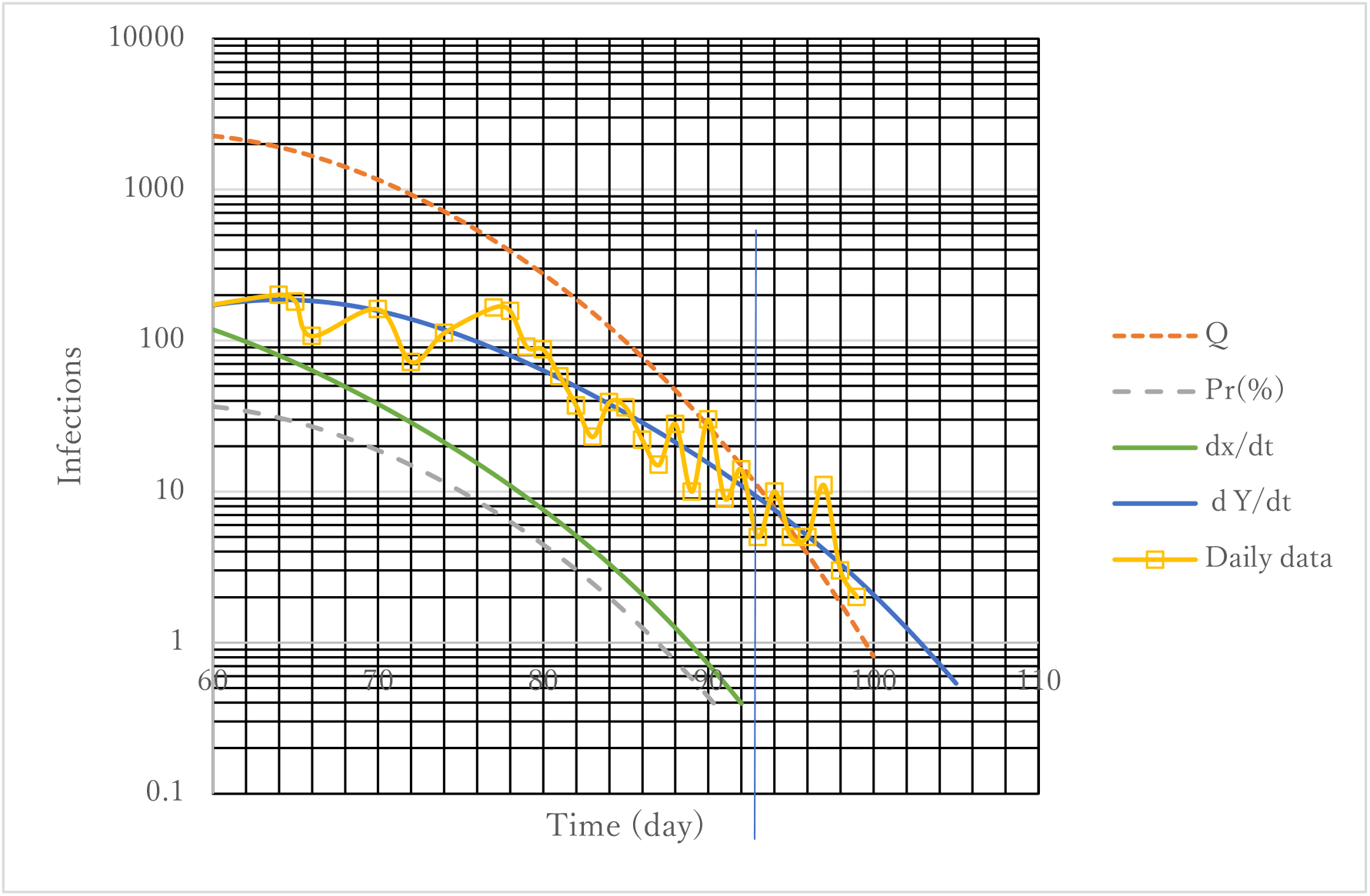
Trends of infectious variables at exit.

### Strategy for Subsequent Epidemics

**Figures 14 and 15** show parametric effects. Calculations with parameter set of *T*=14 and α0 are set for the first epidemic and standard. Smaller α is seen to reduce the second epidemic markedly. A similar effect is expected by reduction of time lag *T* for both cumulative and daily new confirmed cases. Recovery of economic and social activity must accompany infections in a tolerable range. Consequently, reduction in *T* under an increase in α is expected to be the basic policy for the second epidemic.

**Figure 14.**
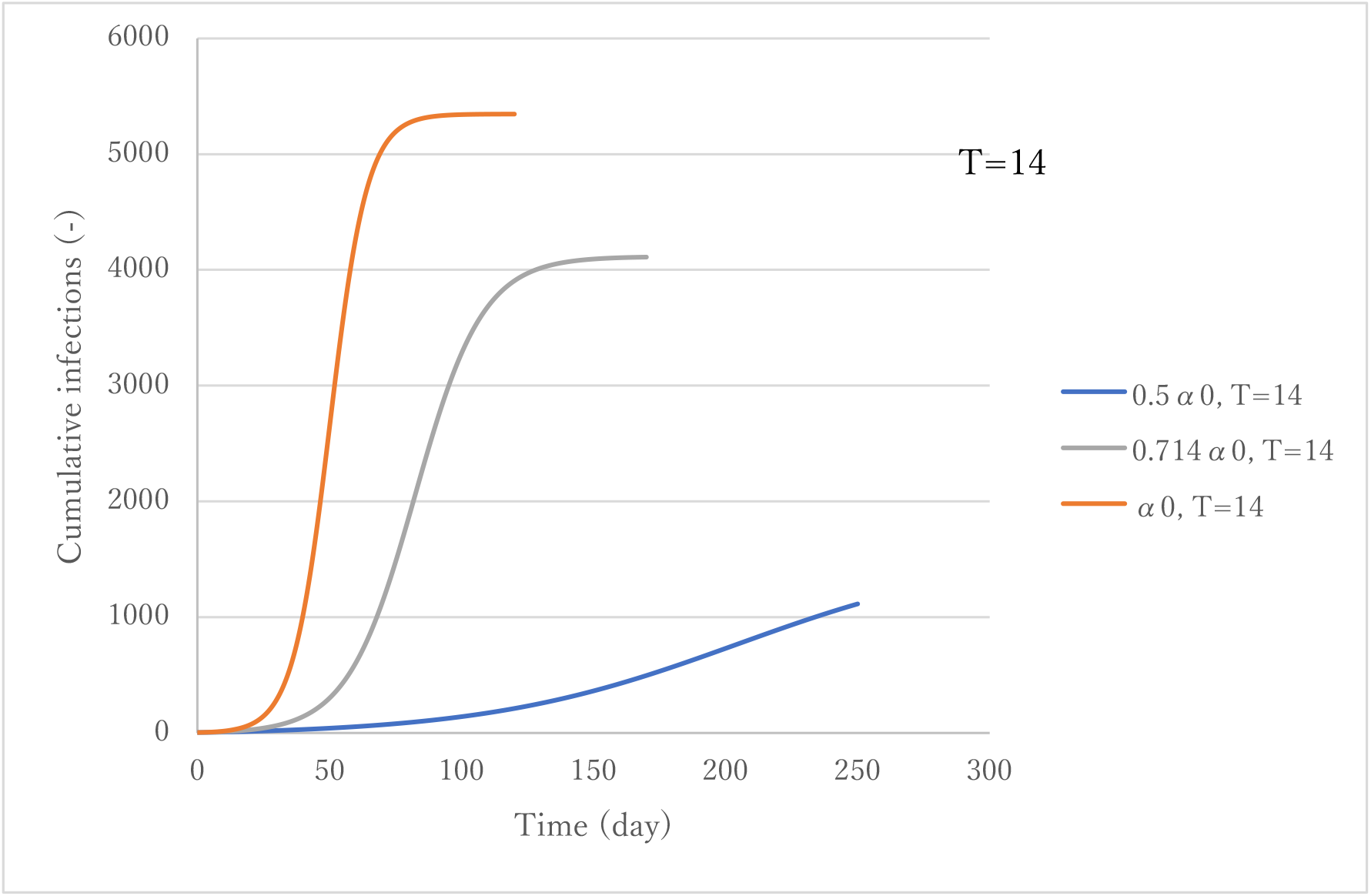
Effect of *α*.

**Figure 15.**
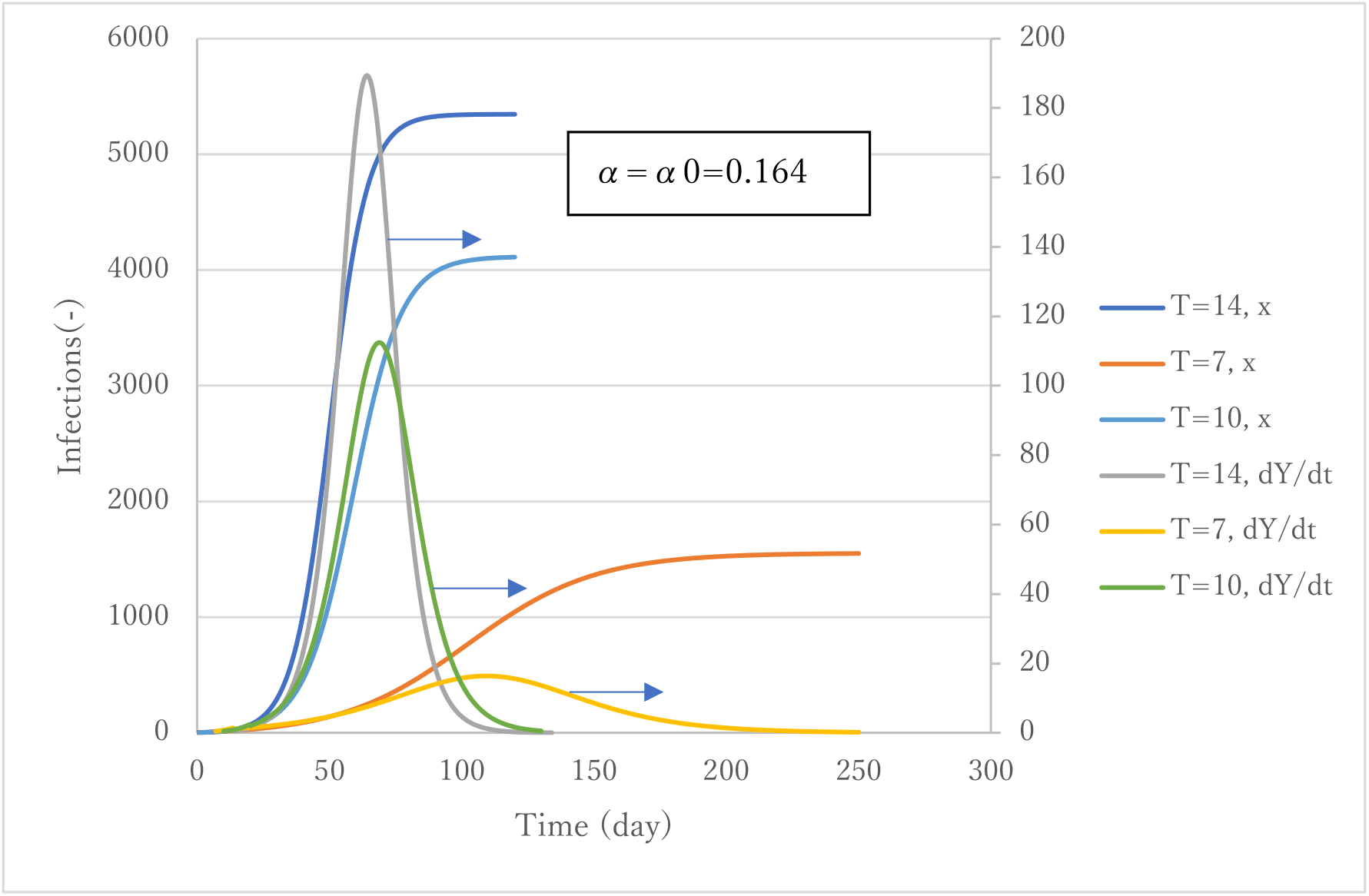
Effect of *T*.

**Figure 16** presents trends of cumulative cases with three (α*T*) values 14α0, 10α0 and 7α0 respectively each of which has two curves. Among them, parameter set of T=14 and α0 is the standard case that simulated the 1^st^ epidemic. It is noteworthy that asymptotic values *x*(∞) of two curves under common α*T* are same although initial increasing rates are different. It is seen the lower the (α*T*), the smaller would become the magnitude of epidemic. From this observation, three sets of parameter combination (α*T, x*(∞)/*M*) being (1.148, 0.251), (1.640, 0.665) and (2.296, 0.862) was obtained. Here, we have derived a final size equation for ATLM in which an attack rate defined as *x*(∞)/*M* is the solution *p* of the following transcendental equation,

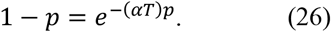

**Figure 16.**
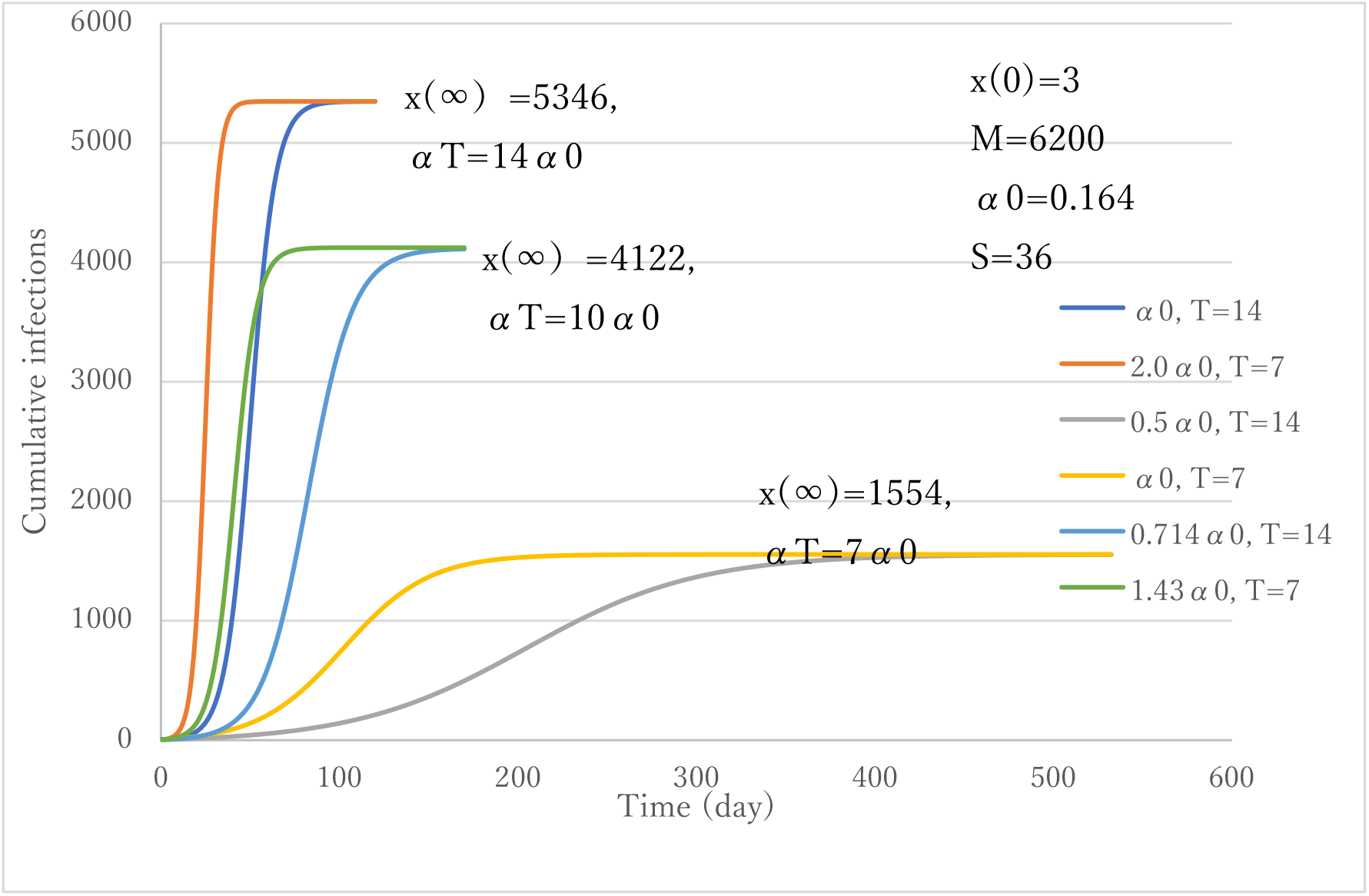
Effect of *αT* on *x*(∞).

Once (α*T*) is known, *x*(∞) would be inferred through Eq.(26). **Figure 17** presents the accuracy of Eq.(26). Excellent agreement with numerical results is obtained. Mathematical proof of Eq.(26) is presented in Appendix-B. In the region (α*T*) <3 strong correlation is observed between *p* and (α*T*) but the attack rate tends to become saturated to unity if (α*T*) exceeds three. It is interesting to note that the parameter (α*T*) dominates not only characteristic of initial stage but also that of endpoint.

**Figure 17.**
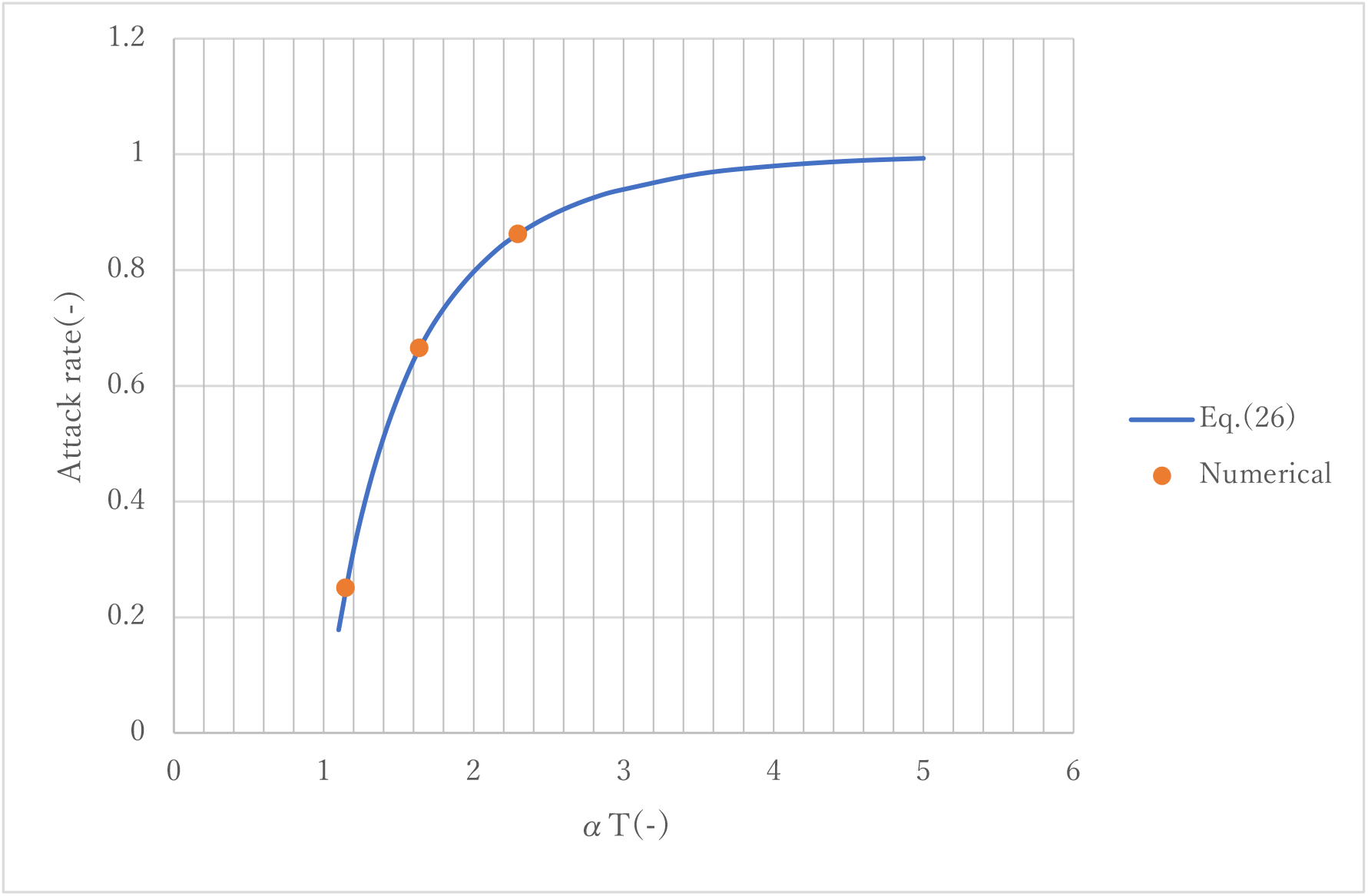
Relationship between attack rate *p* and *αT*.

Among six parameter sets in **Fig. 16** aforementioned, the combination of *T*=7 and *α*=1.43α0 gives an enhanced social activity, but still a smaller magnitude of cumulative infections. To make *T* smaller in practice, faster capture of infected people in the field is necessary. This requires a strong task force to find infectant clusters and to apply rapid testing to the greatest degree possible.

Another point of checking is to prepare necessary number of beds. **Figure 18** exhibits the number of patients under α*T*=10α0. In the case of *T*=7, the maximum number of patients is much the same as in the first epidemic (see **Fig. 4**). Unless *T*=7 is feasible, the second best choice would be *T*=10 with α0.

**Figure 18.**
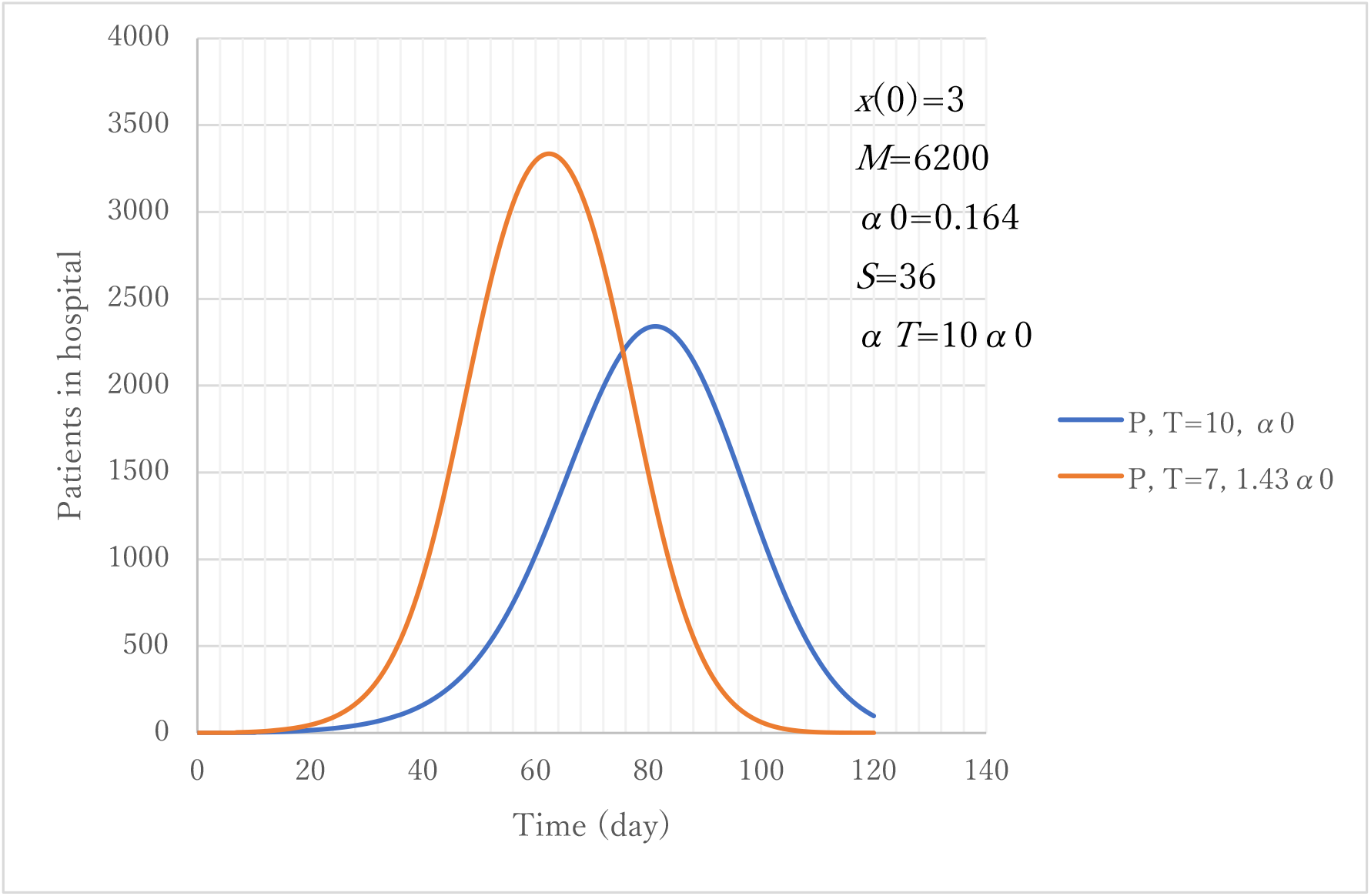
Prediction of the number of hospitalized patients with *T*.

The present model (ATLM) has limitations. First, it is assumed that all new cases spread disease from the time of infection *t* until *t*+*T* isolation in hospitals. In fact, a non-infectant period exists at the nascent stage of incubation. The characteristics of the onset of clinical symptoms are stochastic. Therefore, the modeling of a pre-symptomatic period and statistical process is needed for accurate prediction. Secondly, input parameter *M* (collective population) introduced into ATLM as a parameter should be modelled. Modeling of these might yield insight into clarification of reasons why large differences in the number of cumulative infections exist among countries. These are our tasks for future research. In summary, this study is a first work of detailed quantitative analysis of epidemiological trends of COVID-19 in Tokyo and can be applied easily to COVID-19 trends in other countries.

## CONCLUSIONS

1. An epidemiological model with apparent time delay (ATLM) was developed. It features the capability of implying both the number of patients in hospitals and infectious cases in the field. The model was verified using various epidemic trend data for COVID-19 compiled by the public health authority of metropolitan Tokyo.
2. An alternative form of the standard SIR epidemiological model was obtained and compared with ATLM. The latter underestimated the former by 20% because of effects of isolation of cases.
3. The basic reproductive number was identified as the product (αT) of the present model parameters α and time delay *T*. It was estimated as not less than 2.3 in COVID-19 in Tokyo.
4. Executed intervention to mitigate COVID-19 epidemic spread was assessed in posterior analysis. Its action was found to be too late to be effective. For greater effectiveness, it should have been executed one month earlier in view of the trend of infectious cases in the field.
5. Exit policy made by Japanese government was assessed as proper overall as far as daily new cases were concerned. The date of rescission, however, should have been one week later when remaining infectious cases in the field are below unity.
6. Reduction in *T* with increase in α in model parameters is expected to be the fundamental policy for dealing with a second epidemic to restore social activity while maintaining infections low. Actually, αT=1.64 is an ideal target with *T*=7: the number of beds needed is much the same as in the first epidemic. Unless available technically, *T*=10 would be the second-best choice.

## Data Availability

For this study, we used a publicly available dataset of COVID-19 provided by the public health authority of Tokyo, Japan. These data were collected by public authority announcements. Therefore, ethical approval is not required for the present study.

https://stopcovid19.metro.tokyo.lg.jp/

## DATA AVAILABILITY

We used time-series data of COVID-19 for February 28 through May 23, 2020 in Tokyo. https://stopcovid19.metro.tokyo.lg.jp/

## AUTHOR CONTRIBUTIONS

M. Utamura developed and verified the epidemiological model. M. Koizumi developed the computational method. S. Kirikami engaged with the mathematical aspect of ATLM. All authors have read and agreed to the published version of manuscript.

## ACKNOWLEDGMENT

We are indebted to Mr. Kazushige Tohei for his valuable comments on interpreting numerical results.

## NOMENCLATURE

ATLM: Apparent Time Lag Model

*h*: time step

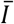: infectious cases

*M*: collective population (in ATLM)

*N*: collective population (in SIR)

*P*: number of patients in hospital (=*Y*-*Z*))

*p*: attack rate (= *x*(∞)/*M*)

*Pr*: positivity-ratio in PCR test (%)

PCR: polymerase chain reaction

*Q*: infectious cases in field (=*x*-*Y*)

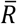: removed cases (recovered/deaths)

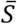: susceptible cases

*s*: variable of Laplace transformation

SEIR: Susceptible, Exposed, Infectious and Removed model

SIR: Susceptible, Infectious and Removed model

*T*: time interval from infection to isolation

*u*: step function

*X*(*s*): Laplace transform of *x*(*t*)

*x*(*t*): cumulative cases

*x*(0): initial value of *x*(*t*)

*x*(∞): magnitude of epidemic

*dx*/*dt*: daily new cases

*Y*: cumulative hospitalized cases (=*x*(*t*-*T*))

*dY*/*dt*: daily hospitalized cases/ daily confirmed cases

*Z*: removed cases (=*x*(*t*-*S*))

**Greek letters**

*α*: transmission rate (in ATLM)

*β*: ibid. (in SIR)

*ε*: share of asymptomatic cases among infectious cases

*τ*: epidemic doubling time

## APPENDIX-A

**Derivation of Eq. (19)**

Under the condition that 1≫*X*/*M*, approximate expression of Eq. (17) is given

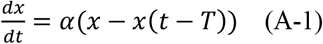

Let *X*(s) be Laplace transformation of *x*(*t*); then Eq. (A-1) would become the following.

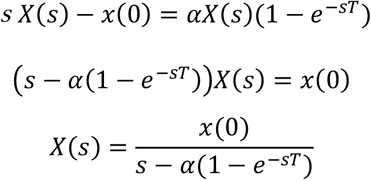

McLaughlin expansion gives

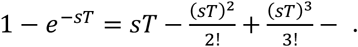

Second-order approximation gives

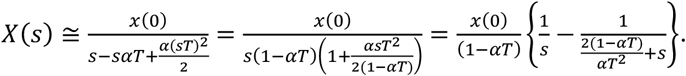

Inverse Laplace transformation of *X*(*s*) gives the following.

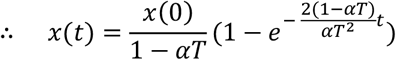

Let *R*0=*αT*, we obtain the expressions shown below.

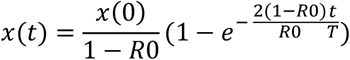

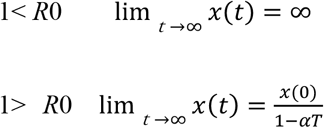

Limitation of *x*(*t*) at *R*0=1 gives the equation below.

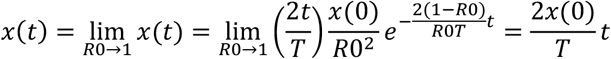

## Appendix B

**Derivation of Equation (26)**

Governing equation of ATLM:

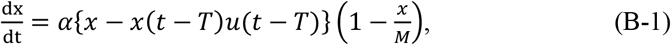

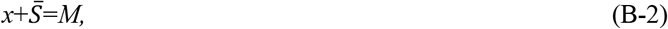

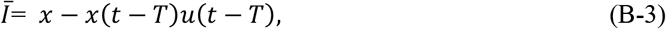

where 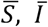 and M are susceptible, infectious and collective number of peoples. Equation (B-1) can be rewritten as

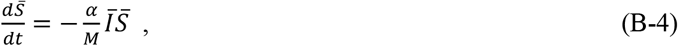

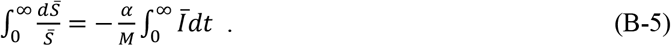

Laplace transformation gives 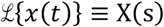, 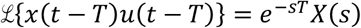. Further expansion gives

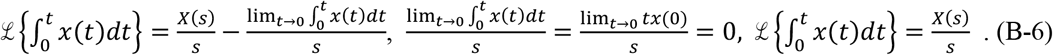

From definition *x*(0−*T*)*u*(0−*T*)=0, then

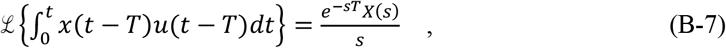

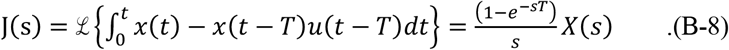

From final value theorem

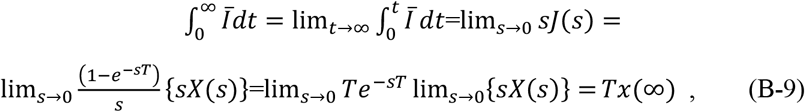

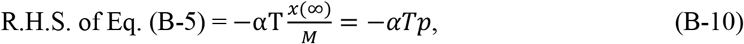

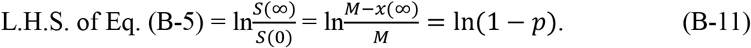

where *p* is the attack rate and is defined by 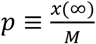. From Eqs. (B-10) and (B-11), we have Eq.(26) as

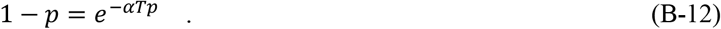

